# The anatomy of regression-to-the-mean in simulated epilepsy trials

**DOI:** 10.64898/2026.07.27.26359051

**Authors:** Daniel M. Goldenholz, Rohan Bhansali, Ted J. Kaptchuk, Brandon Westover

**Affiliations:** Department of Neurology, Beth Israel Deaconess Medical Center, Boston, Massachusetts, USA; Harvard Medical School, Boston, Massachusetts, USA; Program in Placebo Studies, Beth Israel Deaconess Medical Center, Boston, Massachusetts, USA; Department of Neurology, Stanford University, Stanford, California, USA

**Keywords:** epilepsy, regression to the mean, placebo response, seizure diary, clinical trials, seizure detection

## Abstract

Regression to the mean (RTM) can inflate apparent placebo response in epilepsy trials, but its mechanisms are often conflated. Using CHOCOLATES, we simulated 1,000,000 patients with 36 months of daily seizure counts and simulated placebo trials: 2-month baselines followed by 3-month test periods without treatment effects. Transient worsening (RTM type 1), stricter eligibility thresholds (RTM type 2), reduced sensitivity, and false alarms (RTM type 3) each increased RTM and apparent response. These findings show that placebo-arm improvement can arise from temporary illness, natural variability, measurement error, or mixtures thereof, informing epilepsy trial design and endpoint interpretation.

## Introduction

Epilepsy randomized controlled trials (RCTs) commonly enroll people only after a high-seizure baseline period and then judge outcome as change from that baseline. This design is clinically intuitive, but it makes trials vulnerable to apparent improvement that would have happened even without treatment.^1–7^ One important contributor is regression to the mean (RTM), in which a high baseline value is followed by a later value closer to the person’s usual level.^1–3^

The phrase RTM can hide several different causes. RTM type 1 occurs when a person is temporarily more sick than usual: a transient illness, stressor, or other short-lived state raises seizure frequency during baseline and then resolves. RTM type 2 occurs from natural variability alone: eligibility rules select high baseline months from a fluctuating series, even when nothing about the person has changed. RTM type 3 occurs when measurement error changes the apparent seizure-count series, such as missed seizures, false alarms, or diary errors.^8–12^ These mechanisms can coexist, but separating them helps trialists think about prevention.

We used one common simulated cohort to ask whether small, isolated changes could recreate all three RTM subtypes in an epilepsy RCT-like design. We therefore used a realistic seizure-diary simulation to isolate how transient clinical worsening, eligibility selection, and measurement error can each generate placebo-arm seizure reduction in the absence of treatment.

## Methods

We generated 1,000,000 synthetic patients with CHOCOLATES, an open-source simulator designed to reproduce key statistical features of seizure diaries, including heterogeneous long-term rates, clustering, cycles, and the relationship between seizure frequency and variability.^8^ Each patient had 36 months of daily seizure counts. The same underlying cohort was used for every condition.

Eligibility was assessed in rolling 2-month baseline windows, beginning at month 0 and ending at month 31 so that three following test months remained. The first eligible window was used. The primary eligibility rule matched prior focal epilepsy trial criteria: mean baseline seizure frequency at least 4/month, at least 3 seizures in each baseline month, and no baseline seizure-free interval longer than 25 days.^1,13,14^ Patients without any eligible window were not “recruited” and therefore excluded from RTM and MPC summaries but included in eligibility-yield counts. Eligibility and RTM reference rates were recalculated after each condition transform, so each analysis used the observed diary that would have been available under that condition. This simulation used no human participants, human specimens, or identifiable health data.

The baseline condition used a perfect diary: 100% sensitivity and zero false alarms. For RTM type 1, one random consecutive 2-month interval per patient was replaced by a newly simulated CHOCOLATES segment using a newly sampled mean seizure frequency, after which the original seizure frequency resumed. This was done to simulate the temporary effects of something unusual, such as illness. For RTM type 2, only the eligibility threshold changed, from 4 to 8 seizures/month, to create additional pressure for high frequency patients to be identified. For RTM type 3, we separately reduced detector sensitivity to 90%, 80%, and 70% with zero false alarms, and added false alarms at 1/month, 1/week, and 1/day with 100% sensitivity.

Expected false alarms were subtracted for the false-alarm analyses (eligibility and percentage change)^12^.

There was no active treatment effect. RTM was counted when the baseline rate was above the patient’s 36-month observed reference rate and the test-period rate moved closer to that reference than baseline^1^. Placebo response was summarized as MPC, the median percentage change from baseline to test. No hypothesis tests were performed because the analysis was a descriptive simulation rather than a randomized group comparison. Source code is open source at https://github.com/GoldenholzLab/anatomy_of_RTM.git.

## Results

Under the perfect-diary baseline condition, 649,098 of 1,000,000 patients (64.9%) became eligible. Among eligible patients, 53.9% met the RTM definition and MPC was 26.7% (Figure 2). Thus, even without treatment, the usual baseline-versus-test design produced substantial apparent improvement.

**Figure 1.**
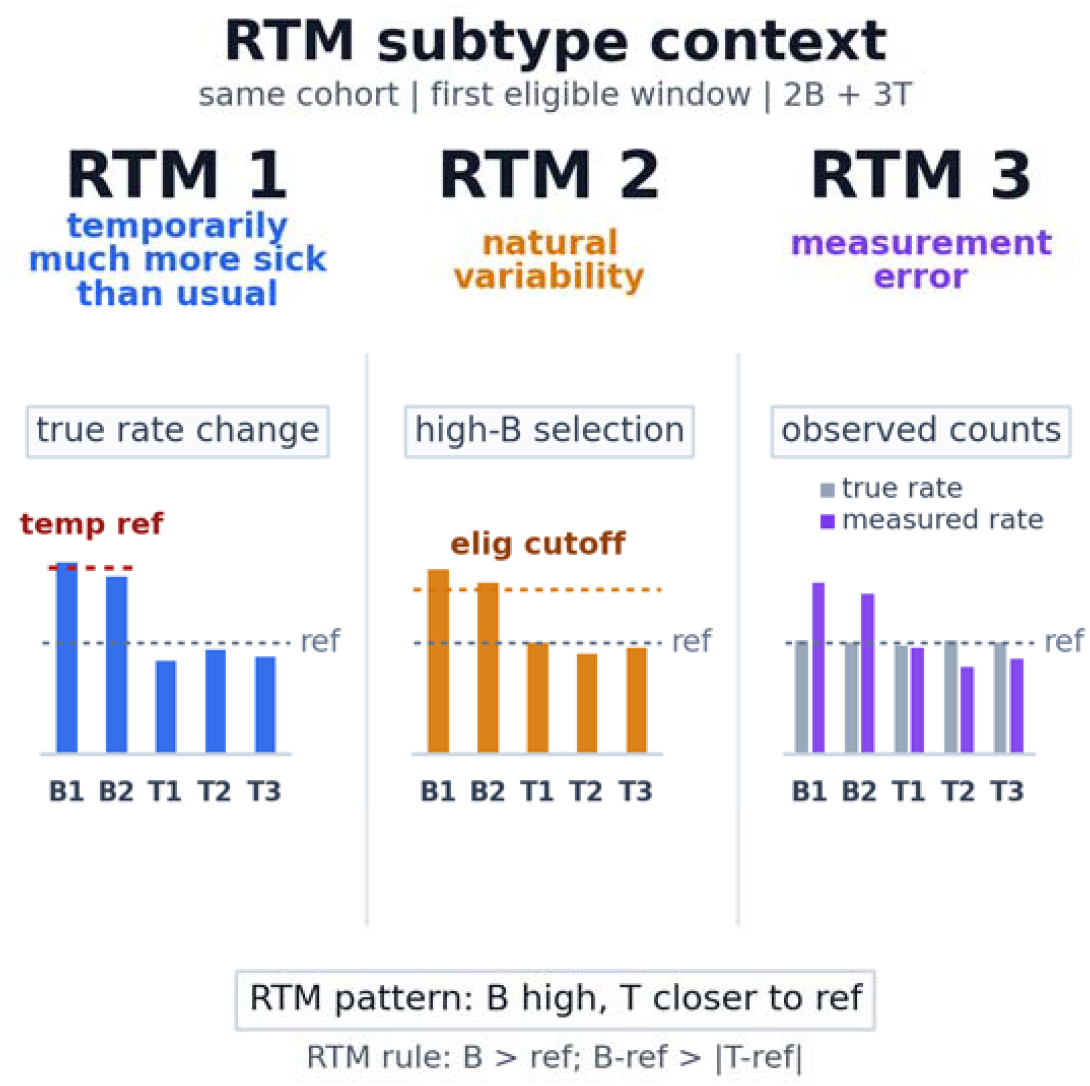
Conceptual overview of the three RTM subtypes. RTM type 1 reflects temporary worsening, represented here as a short-lived higher reference rate during baseline. RTM type 2 reflects natural variability plus eligibility selection, which preferentially selects high baseline months. RTM type 3 reflects measurement error, in which the measured rate differs from the true rate because of missed events, false alarms, or other count errors. B1 and B2 denote the 2-month baseline; T1-T3 denote the 3-month test period; ref denotes the long-term reference rate.

**Figure 2.**
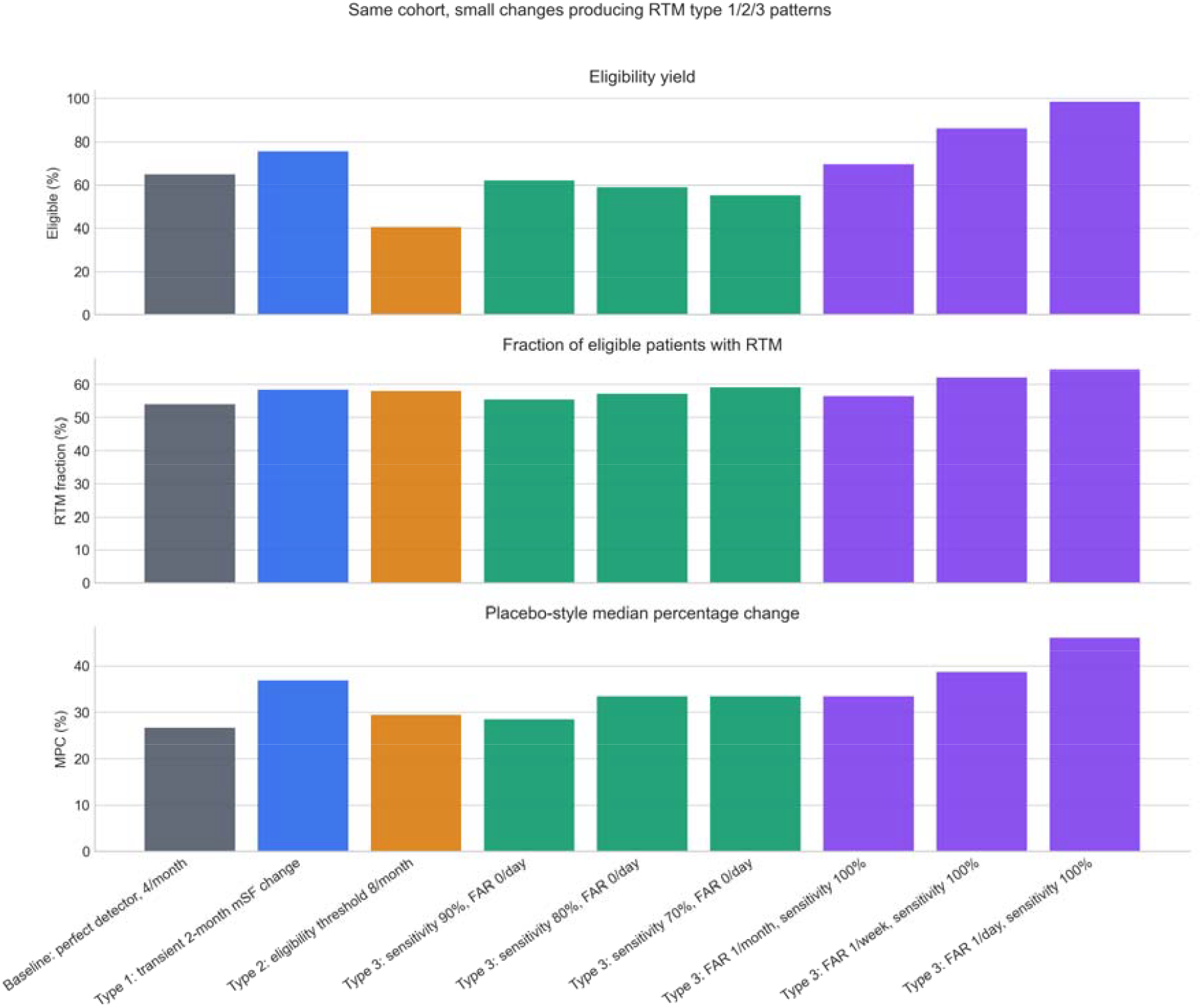
One-cohort RTM type 1/2/3 simulation results. All conditions used the same 1,000,000-patient CHOCOLATES cohort with 36 months of daily data and first eligible baseline selection. Panels show eligibility yield, fraction of eligible patients with RTM, and placebo median percentage change (MPC). The baseline condition used a perfect detector and the 4 seizures/month eligibility threshold. RTM, regression to the mean; MPC, median percentage change; FAR, false alarm rate.

The RTM type 1 condition sometimes made patients temporarily much more sick than usual in one randomly selected 2-month interval. Eligibility increased to 75.6%, RTM increased to 58.4%, and MPC increased to 36.8%. The effect depended strongly on where the transient worsening occurred. When the transient interval overlapped the selected baseline, RTM was 95.1% and MPC was 92.6%. When it overlapped neither baseline nor test, RTM and MPC were closer to the baseline condition (51.1% and 25.9%).

The RTM type 2 condition changed only one eligibility number, from 4 to 8 seizures/month. This made eligibility stricter, reducing the eligible group to 40.4% of the cohort, but it also selected more extreme baseline periods. RTM rose to 58.0% and MPC rose to 29.4%. This demonstrates how natural variability alone can change the apparent placebo response when entry rules are adjusted.

The RTM type 3 conditions altered measurement. With false alarms fixed at zero, reducing sensitivity from 100% to 70% reduced eligibility from 64.9% to 55.2%, increased RTM from 53.9% to 59.1%, and increased MPC from 26.7% to 33.3%. With sensitivity fixed at 100%, increasing false alarms from zero to 1/day increased eligibility to 98.5%, RTM to 64.4%, and MPC to 46.0%, despite expected false-alarm correction.

## Discussion

This one-cohort simulation supports a simple message: RTM in epilepsy trials is not one mechanism. Temporary worsening, natural seizure variability, and measurement error can each make baseline look worse than the later test period. All three produce the same trial-facing result - apparent placebo improvement - but their practical implications differ.

RTM type 1 is clinically intuitive. If a participant enters a trial during a transient bad period, improvement after enrollment may reflect recovery from that temporary state. RTM type 2 is more statistical: the eligibility rule preferentially captures high points from an otherwise fluctuating diary. RTM type 3 is increasingly relevant as trials consider device-derived seizure counts. Automated detection can reduce under-reporting, but sensitivity, false alarm rate, and correction rules become part of the endpoint itself.^9–12^

The results also show why placebo response should not be interpreted as a single psychological placebo effect. In these simulations there was no expectation, blinding response, medication ritual, or active therapy. The observed response arose from how patients were selected and how seizures were measured. This does not necessarily mean psychological placebo effects are absent in real trials.^4,15^ These findings do however support experimental findings that suggest that placebo responses in situations measuring objective outcomes are primarily, if not exclusively, regression to the mean^16^ whereas purely subjective reported outcomes involve neuropsychological processes^17,18^. In general, this finding means that baseline selection and measurement should be modeled before assigning biological or behavioral meaning to placebo-arm improvement.

These findings have practical design implications. Eligibility criteria should be simulated before trials begin, especially when changing the seizure-frequency threshold or baseline duration.

Device-based trials should prespecify how sensitivity, false alarms, false-alarm correction, missing wear time, and seizure-type-specific detection enter eligibility and endpoints. When feasible, sensitivity analyses should separate true-rate assumptions from measurement assumptions.

Limitations should be noted. CHOCOLATES is realistic but synthetic, and this analysis used synthetic participants only. The RTM type 1 intervention was a simplified 2-month rate change, not a data-driven clinical illness model. Detector sensitivity and false alarms were uniform across people and time, whereas real devices may vary by patient, seizure type, state, and adherence.

The false-alarm correction was idealized. Finally, these simulations describe apparent placebo response, not drug-placebo separation in a randomized active-treatment trial.

In summary, a single simulated epilepsy cohort can express RTM type 1, type 2, or type 3 when only one design or measurement feature is changed. Recognizing these subtypes may make placebo response less mysterious and improve how epilepsy RCTs are designed, interpreted, and simulated.

## Data Availability

The open-source code for this study is available at https://github.com/GoldenholzLab/anatomy_of_RTM.git. The analysis uses CHOCOLATES, available at https://github.com/GoldenholzLab/CHOCOLATE.git.

## Acknowledgments and funding

DMG was funded by the National Institutes of Health (NIH) K23NS124656, 1R21NS142800 and the American Board of Psychiatry and Neurology (ABPN). RB was funded by NIH K23NS124656. OpenAI Codex was used to assist with coding and portions of manuscript preparation, however the authors take full responsibility for the entire project including code, images, and manuscript.

## Ethical publication statement

We confirm that we have read the Journal’s position on issues involved in ethical publication and affirm that this report is consistent with those guidelines.

## Author contributions

Daniel M. Goldenholz conceived the analysis, wrote and revised analysis code, interpreted results, and drafted the manuscript. Rohan Bhansali contributed to interpretation and manuscript revision. Ted Kaptchuk contributed to interpretation and manuscript revision. Brandon Westover contributed to interpretation, simulation framing, and manuscript revision.

